# Low-dose calcium supplementation during pregnancy in low and middle-income countries: a cost-effectiveness analysis

**DOI:** 10.1101/2024.11.14.24317327

**Authors:** Happiness P. Saronga, Pratibha Dwarkanath, Hening Cui, Alfa Muhihi, Anura V. Kurpad, V. Sri Poornima, Mary M. Sando, R. Poornima, Cara Yelverton, Honorati M. Masanja, Christopher R. Sudfeld, Andrea B. Pembe, Wafaie W. Fawzi, Nicolas A. Menzies

## Abstract

**Background:** Calcium supplementation during pregnancy can reduce the risk of preeclampsia and preterm birth. Few countries have implemented WHO-recommended high-dose calcium supplementation (1500-2000mg/day), due to adherence and cost concerns. However, low-dose calcium supplementation (one 500mg tablet daily) has recently been shown to be similarly efficacious as high-dose supplementation. We assessed the cost-effectiveness of low-dose calcium supplementation during pregnancy, in low- and middle-income countries (LMICs) with low dietary calcium intake.

**Methods:** Using a mathematical model, we estimated the lifetime health outcomes (cases, deaths and DALYs averted) and costs of low-dose calcium supplementation provided through routine antenatal care to women giving birth in 2024, as compared to no supplementation. We assessed costs (2022 USD) from a health system perspective, including cost-savings from averted care for preeclampsia and preterm birth.

**Findings:** Low-dose calcium supplementation was estimated to prevent 1.3 (95% uncertainty interval: 0.2, 2.6) million preterm births (a 10% (2, 18) reduction), 1.8 (1.0, 2.8) million preeclampsia cases (a 23% (14, 32) reduction), as well as 5.9 (1.3, 12.9) million disability-adjusted life years (DALYs). Intervention costs would be $267 (220, 318) million and produce cost-savings of $56 (26, 86) million, with incremental costs per DALY averted of $90 (38, 389) across all countries, and a return on investment of 19.1 (3.8, 39.5). The intervention was cost-effective in 119 of 129 countries modeled when compared to setting-specific cost-effectiveness thresholds. While there was substantial uncertainty in several inputs, cost-effeciveness conclusions were robust to parameter uncertainty and alternative analytic assumptions.

**Interpretation:** Low-dose calcium supplementation provided during pregnancy is cost-effective for prevention of preeclampsia and preterm birth in most LMICs.

## Funding

Bill & Melinda Gates Foundation.

## Introduction

Hypertensive disorders of pregnancy (HDP) and preterm births are leading global contributors to mortality and morbidity for mothers and infants. HDP—which includes preeclampsia and eclampsia—cause an estimated 14% of all maternal deaths and are major risk factors for preterm birth,^1^ the leading global cause of death among children <5 years.^2^

The World Health Organization (WHO) currently recommends that pregnant women living in areas with low calcium intake receive high-dose calcium supplementation to reduce the risk of preeclampsia.^3^ The WHO-recommended dosing scheme for high-dose calcium supplementation is 1500-2000mg/day, divided into three doses and taken a few hours apart from iron-folic acid (IFA) supplements to reduce potential negative effects on iron absorption. In addition to adherence concerns due to the complexity of the dosing scheme, the costs associated with implementing high-dose calcium supplementation have also impeded scale-up. The additional cost of three-tablet high-dose calcium supplementation is estimated to be US$9-11 per pregnancy, which is substantially more than the current costs of iron-folic acid supplementation (US$1-2 per pregnancy).^3^ As a result of these adherence and cost concerns, few low-and middle-income countries (LMIC) have implemented routine calcium supplementation in pregnancy.

One possible approach for enabling the scale-up of prenatal calcium supplementation is to reduce the number of doses per day, thereby reducing the pill burden and program costs. While trials of low-dose (<1000mg/day) calcium supplementation during pregnancy have shown similar reductions in risks of preeclampsia and preterm birth as high-dose calcium supplementation;^4^ until recently there had not been trials that directly compared low-dose with high-dose calcium supplementation in pregnancy. Two independent double-blind, parallel-group randomized non-inferiority trials of low-dose (500 mg/day) vs. high-dose (1500 mg/day) calcium supplementation in pregnancy have now been conducted in Tanzania and India.^5^ These trials showed that low-dose supplementation was non-inferior to high-dose supplementation with respect to the risk of preeclampsia in both countries, and non-inferior with respect to preterm birth in India. Therefore, low-dose and high-dose calcium supplementation appear to be similarly efficacious, consistent with systematic reviews reporting similar magnitudes of impact.^4^

While past studies have evaluated the cost-effectiveness and return on investment of high-dose calcium supplementation,^6–9^ these studies have reported divergent results. Additional cost-effectiveness evidence for calcium supplementation has been identified as a key research priority by a taskforce convened to assess the evidence base around global calcium deficiency.^10^ In particular, there is limited evidence on the cost-effectiveness of low-dose calcium supplementation. No prior study has assessed the cost-effectiveness, return-on-investment and cost-savings possible with low-dose calcium supplementation in low- and middle-income settings, where the majority of preeclampsia cases and pre-terms births occur. In this study, we estimated the potential health impact, cost-effectiveness, and resource needs for introducing low-dose calcium supplementation in pregnancy to prevent preeclampsia and preterm birth. We estimated results for low- and middle-income countries with low dietary calcium intake, considering a range of assumptions for coverage, adherence, supplementation cost, sources of evidence for intervention impact, and other factors determining intervention impact and costs.

## Methods

We constructed a mathematical model estimating pregnancy outcomes in each LMIC for the 2024 birth cohort. Using this model, we simulated the reductions in preeclampsia and preterm birth that would be produced by a low-dose calcium supplementation intervention delivered through routine antenatal care (ANC). We calculated disability-adjusted life years (DALYs) to quantify health benefits, and calculated intervention costs as well as cost-savings due to averted care for pregnancy complications. Using these outcomes, we estimated incremental cost-effectiveness ratios (ICERs) and compared these to country-specific cost-effectiveness thresholds to summarize intervention cost-effectiveness in each country. We estimated budgetary requirements for intervention introduction, as well as the net monetary benefit, return on investment, and cost-savings of this intervention compared to high-dose calcium supplementation.

### Intervention scenario

We constructed a base-case scenario that assumed no calcium supplementation during pregnancy, as many countries do not currently provide calcium supplementation during pregnancy despite this intervention being included in WHO recommendations.^3^ We compared this base-case to an intervention scenario assuming that women would be provided with low-dose calcium supplementation (one pill to be consumed daily containing 500mg elemental calcium) from 20 weeks of gestational age until delivery. We assumed the intervention would only be offered in countries with low dietary calcium among women of reproductive age (defined as having >25% of individuals with calcium intake of <800mg/day), would only be received by women with consistent ANC attendance (defined as attending 4+ ANC visits during pregnancy), and that intervention adherence in routine programmatic settings would be lower than observed in clinical trials. Base-case and intervention scenarios were evaluated for the 2024 birth cohort in each LMIC based on World Bank criteria (per capita gross national income less than US$13,846 in 2022).

### Calculation of birth outcomes

#### Base-case scenario

Numbers of births for each country were based on UN Population Division projections for 2024. The proportion of preterm births was based on a recent study estimating prematurity by country in 2020.^11^ The proportion of births with preeclampsia was calculated by taking estimated incidence of maternal hypertensive disorders in 2019 from the Global Burden of Disease Study, scaling these values by the fraction with preeclampsia, and dividing by annual numbers of live births.^12,13^ Numbers of preterm births and preeclampsia cases under the base-case scenario were calculated based on these inputs.

#### Intervention scenario

To identify countries with low dietary calcium in women of reproductive age we estimated country-level distributions of calcium intake within each age group and sex employing a published approach,^14^ and adjusted these distributions to match average calcium intake estimates for each country.^15^ Based on these distributions we estimated the proportion of women aged 15-49 years old with <800mg/day calcium intake. If this proportion was >25% the country was assumed to introduce the intervention. The proportion of women attending 4+ ANC visits was derived from country data collated by WHO. We further assumed that intervention adherence in routine programmatic settings would be lower than observed in clinical trials, and estimated adherence based on reported data for routine IFA supplementation, operationalized as the proportion of women reporting taking >90 iron-folate pills during their last pregnancy within the last 3 years, among all women reporting 4+ ANC visits and receiving any iron-folate supplementation (*high-adherence*). This ratio was estimated from recent Demographic and Health Survey data. Intervention efficacy was based on a published meta-analyses of high-dose (>1000mg/day) calcium supplementation compared to placebo for prevention of preterm birth (risk ratio (RR) 0.76 (95% confidence interval (CI): 0.60, 0.97)) and preeclampsia (RR 0.45 (95% CI: 0.31, 0.65)),^4^ following recent trial results showing low-dose and high-dose calcium supplementation during pregnancy to be similarly efficacious.^5^ As these RRs were obtained under trial conditions we assumed they would only apply to high-adherence intervention recipients. To be conservative, we assumed that RRs would be 1.0 (i.e., no preventive effect) for low-adherence recipients. Reductions in the number of cases of preterm birth and preeclampsia were calculated for each country based on these inputs, comparing the intervention scenario to the base-case. Analytic equations are provided in the Supplement.

### Calculation of summary health outcomes

We estimated the DALYs averted by the intervention to quantify health benefits over the lifetime of affected individuals, combining fatal (Years of Life Lost (YLLs)) and non-fatal (Years Lived with Disability (YLDs)) health losses experienced by mothers and infants. The number of YLLs and YLDs per case of preterm birth and preeclampsia were estimated from results reported by the Global Burden of Disease Study.^12^ These ratios were calculated for each country, and applied to the estimated numbers of preterm births and preeclampsia cases averted. DALYs estimated were summarized by condition (preterm birth, preeclampsia), age group (infants, mothers), form of health loss (fatal, non-fatal) and country. We also calculated deaths averted by the intervention, based on estimates of the deaths per case of preterm birth and preeclampsia.^12^

### Cost estimation

Costs were estimated from a health system perspective. The unit cost per 500 mg calcium tablet was assumed to be US$0.02, based on current global price levels.^16^ We assumed 20 weeks of supplementation with one pill daily (140 pills total). We applied a 6% wastage rate and a 13% mark-up for supply chain costs (total cost per pregnancy $3.37).^17,18^ We assumed the intervention would be delivered through routine antenatal care, with no additional clinic visits required.

To estimate the reduction in healthcare costs due to reduced needs for preterm birth care, we estimated the incremental costs per preterm birth and applied this to number of preterm births averted for each country. Incremental costs per preterm birth, and per preterm birth resulting in infant death (versus term birth), were estimated from a multi-country costing study.^19^ A similar approach was taken to calculate the reduction in health care costs due to reduced needs for preeclampsia care, with the incremental costs per preeclampsia case estimated from the cost difference between *high-risk* and *non-high-risk* pregnancies,^19^ and applied to the number of averted preeclampsia cases for each country. To adjust unit costs to different countries we applied WHO-CHOICE estimates of the elasticity of healthcare costs to country income (0.87 (95% CI: 0.83, 0.91) increase in log unit costs for every 1 unit increase in log per capita GDP).^20^ Cost inputs were inflated to 2022 price levels using the GDP deflator and reported in 2022 US dollars.

### Cost-effectiveness analysis

We estimated ICERs as the incremental cost per DALY averted, dividing mean incremental costs by mean incremental benefits, for the intervention scenario compared to the base-case. For each country we compared the ICER to published estimates of the opportunity cost of healthcare spending,^21^ to report whether the intervention would be cost-effective in each country. These thresholds have been proposed as a realistic estimate of the marginal health benefits produced by healthcare spending, and provide a proxy of what is given up when additional resources are devoted to a new intervention in the absence of additional funding.

### Additional economic outcomes

We estimated several additional outcomes describing the value of low-dose calcium supplementation: (i) costs per adverse pregnancy outcome (sum of preterm births and preeclampsia cases) averted; (ii) net monetary benefits (NMB), calculated as the monetary value of health benefits minus net costs; (iii) return on investment (ROI), calculated as the ratio of NMB to intervention costs; and (iv) cost savings of the intervention relative to high-dose calcium supplementation of 1500mg/day (3 calcium tablets).^16^ While cost and health outcomes are reported undiscounted, we applied a 3% discount rate when calculating ICERs, NMB, and ROI.^22^ Analytic equations are provided in the Supplement.

### Imputation of missing values

We imputed missing values for several analytic inputs (details in Supplement). Figure S1 describes availability of inputs for each country.

### Uncertainty and sensitivity analyses

A Monte Carlo simulation approach was used to generate 95% uncertainty intervals. To do so, we specified probability distributions representing uncertainty in each model parameter (Table S1), and re-estimated results for 1000 Latin hypercube parameter samples. For parameters that varied between countries, we assumed uncertainty was correlated across counties (rho = 1.0). We calculated 95% uncertainty intervals as the 2.5^th^ and 97.5^th^ percentiles of the simulation results for each outcome. Analyses were conducted in R v4.0.2.

We conducted one-way deterministic sensitivity analyses quantifying the sensitivity of results to individual parameter changes, varying each parameter between its lower and upper bounds while holding other parameters at their mean value. We estimated these results for the ICER and for the NMB of the intervention across all LMICs.

Several additional sensitivity analyses were conducted to test the robustness of results to changes in key assumptions. First, we calculated health impact results for two additional intervention scenarios: one that assumed high adherence for all women receiving the intervention (*high adherence* scenario), and one that assumed full coverage for the intervention (i.e., not limited by ANC coverage) as well as high adherence (*full coverage and high adherence* scenario). As achieving these scenarios would require additional public health investments beyond those included in our analysis, we did not calculate economic outcomes for them.

Second, we recalculated results assuming low-adherence women would receive 50% of the preventive effect of calcium supplementation (vs. 0% assumed in the main analysis). Third, we recalculated results using published meta-analysis results for low-dose calcium (RR for preterm birth: 0.83 (95% CI: 0.34, 2.03); RR for preeclampsia: 0.38 (95% CI: 0.28, 0.52)).^4^ Fourth, we recalculated economic outcomes excluding the cost-savings from averted care for pregnancy complications, providing a conservative estimate of cost-effectiveness. Finally, we re-estimated economic outcomes for calcium unit costs of US$0.015 and US$0.03, as might be possible with volume-based price reductions, or with price inflation respectively.

### Role of the funder

The funder played no role in study design, implementation, or reporting.

## Results

Across all LMICs, we estimated that 129 of 133 countries have low dietary calcium in women of reproductive age, representing >99% of all LMIC births. Based on published studies we estimated there would be 121 (95% uncertainty interval: 108, 136) million births, 12 (7, 18) million preterm births (representing 10% (6, 15) of all births) and 8 (5, 11) preeclampsia cases (representing 7% (5, 9) of all births) in these countries in 2024.

### Health impact of supplementation

We modelled all LMIC with low dietary calcium in women of reproductive age as introducing the intervention. Based on estimated ANC coverage in each country we assumed that 79 (66, 93) million women would receive the intervention. Table 1 presents health impact results for low-dose calcium supplementation, aggregated by WHO region and income level. Across all modelled countries we estimated the intervention would prevent 1.3 (0.2, 2.7) million preterm births and 1.8 (1.0, 2.8) million preeclampsia cases, representing 10% (2, 18) and 23% (14, 32) of total LMIC preterm births and preeclampsia cases, respectively. As a consequence of these reductions, we estimated there would be 65 (15, 137) thousand fewer deaths and 5.9 (1.3, 12.9) million fewer DALYs attributable to these health conditions in 2024. Across all countries, the large majority of DALYs averted were due to reductions in preterm birth (92% (68, 97), vs. reductions in preeclampsia), were accrued by infants (88% (65, 94), vs. mothers), and resulted from reductions in premature death (94% (91, 95), vs. reductions in non-fatal conditions). Table S2 reports global and regional estimates of deaths and DALYs averted by condition, age group, and form of health loss. Across individual countries India was estimated to have the greatest absolute health gain, representing 1514 (306, 3084) thousand DALYs (26% (22, 30) of the total, Table S3).

**Table 1:** Health effects estimated for the low-dose calcium supplementation intervention in 2024, compared to no intervention. DALY = disability-adjusted life year, ‘000s’ indicates thousands. Values in parentheses represent equal-tailed 95% uncertainty intervals. Results exclude high income countries, countries not assessed as having low dietary calcium among women of reproductive age, or countries with insufficient data.

| Country grouping | Preterm births averted (000s) | Preterm births averted (% of base-case) | Preeclampsia cases averted (000s) | Preeclampsia cases averted (% of base-case) | Infant and maternal deaths averted (000s) | DALYs averted (000s) |
| --- | --- | --- | --- | --- | --- | --- |
| All low and middle-income countries | 1259<br>(209, 2681) | 10%<br>(2, 18) | 1835<br>(1036, 2836) | 23%<br>(14, 32) | 64.9<br>(15.4, 136.7) | 5930<br>(1301, 12877) |
| World Region |  |  |  |  |  |  |
| *Africa* | 339<br>(51, 792) | 8%<br>(1, 14) | 674<br>(377, 1053) | 18%<br>(11, 26) | 21.8<br>(4.6, 49.9) | 1931<br>(389, 4540) |
| *Americas* | 126<br>(23, 238) | 16%<br>(3, 29) | 173<br>(101, 261) | 38%<br>(24, 50) | 4.8<br>(1.3, 8.9) | 449<br>(113, 841) |
| *Eastern Mediterranean* | 181<br>(27, 430) | 9%<br>(2, 16) | 204<br>(105, 339) | 21%<br>(12, 31) | 10.5<br>(2.7, 23.5) | 916<br>(211, 2112) |
| *Europe* | 60<br>(11, 112) | 16%<br>(3, 28) | 91<br>(49, 144) | 37%<br>(24, 49) | 1.8<br>(0.4, 3.5) | 182<br>(38, 354) |
| *South-East Asia* | 379<br>(62, 786) | 9%<br>(2, 17) | 443<br>(252, 674) | 22%<br>(14, 31) | 20.6<br>(4.9, 41.0) | 1914<br>(414, 3921) |
| *Western Pacific* | 174<br>(30, 351) | 16%<br>(3, 28) | 250<br>(144, 381) | 36%<br>(23, 48) | 5.4<br>(1.2, 11.1) | 538<br>(114, 1112) |
| Income Level |  |  |  |  |  |  |
| *Low income* | 171<br>(24, 427) | 6%<br>(1, 12) | 294<br>(151, 492) | 15%<br>(8, 23) | 10.4<br>(2.2, 25.0) | 912<br>(177, 2239) |
| *Lower middle-income* | 716<br>(116, 1559) | 9%<br>(2, 17) | 1008<br>(568, 1557) | 22%<br>(14, 31) | 41.4<br>(9.6, 87.8) | 3752<br>(806, 8195) |
| *Upper middle-income* | 372<br>(65, 718) | 16%<br>(3, 28) | 533<br>(307, 813) | 36%<br>(23, 48) | 13.1<br>(3.3, 25.2) | 1267<br>(295, 2472) |

### Cost-effectiveness

Implementing the intervention was estimated to cost US$267 (220, 318) million across countries introducing the intervention, while cost-savings from averted maternity care were estimated as $56 (26, 86) million, for a total incremental cost of $211 (160, 267) million. The incremental cost per DALY averted was $90 (38, 389) when aggregated across all countries, ranging between $72 and $140 across WHO regions. Across all countries, the cost per adverse pregnancy outcome averted was $75 (38, 144). Table 2 reports estimated costs, cost-effectiveness, and other economic outcomes of low-dose calcium supplementation, aggregated by WHO region and country income level. Across individual countries, the intervention was found to be cost-effective (cost per DALY averted below the cost-effectiveness threshold for each setting) in 119 of 129 countries with low-dietary calcium in women of reproductive age. Table S4 gives country-specific results, in which ICERs ranged from $37 to $412 per DALY averted. Figure 1 shows how cost-effectiveness ratios compare to per-capita GDP for each WHO region and income group.

**Table 2:** Intervention costs, cost savings, and cost-effectiveness for the low-dose calcium supplementation intervention in 2024, compared to no intervention. USD = 2022 US dollars, DALY = disability-adjusted life year, ‘mil.’ indicates millions. Values in parentheses represent equal-tailed 95% uncertainty intervals. Negative values for net monetary benefit or return on investment indicates that the monetary value of health gains is less than incremental costs (i.e., intervention not cost-effective at willingness to pay threshold used for analysis). Results exclude high income countries, countries not assessed as having low dietary calcium among women of reproductive age, or countries with insufficient data.

| Country grouping | Intervention costs (USD mil.) | Cost savings from averted healthcare (USD mil.) | Cost per adverse pregnancy outcome averted (USD) | Incremental cost per DALY averted (USD) | Net monetary benefits (USD mil.) | Return on investment | Cost savings vs. high-dose supplementation (USD, mil.) |
| --- | --- | --- | --- | --- | --- | --- | --- |
| All low and middle-income countries* | 267<br>(220, 318) | 56<br>(26, 86) | 75<br>(38, 144) | 90<br>(38, 389) | 5063<br>(1017, 10417) | 19.1<br>(3.8, 39.5) | 534<br>(440, 637) |
| World Region |  |  |  |  |  |  |  |
| *Africa* | 75<br>(61, 91) | 9<br>(4, 14) | 71<br>(38, 129) | 92<br>(38, 407) | 623<br>(76, 1493) | 8.3<br>(1.0, 19.9) | 150<br>(122, 182) |
| *Americas* | 26<br>(22, 32) | 11<br>(5, 17) | 58<br>(24, 124) | 84<br>(31, 373) | 1156<br>(274, 2215) | 44.0<br>(10.5, 84.7) | 53<br>(43, 63) |
| *Eastern Mediterranean* | 36<br>(26, 47) | 5<br>(2, 8) | 93<br>(43, 184) | 90<br>(38, 357) | 311<br>(47, 735) | 8.6<br>(1.3, 20.3) | 73<br>(52, 94) |
| *Europe* | 15<br>(11, 18) | 6<br>(3, 9) | 65<br>(27, 138) | 113<br>(42, 534) | 463<br>(96, 905) | 31.7<br>(6.6, 61.9) | 29<br>(23, 36) |
| *South-East Asia* | 66<br>(57, 77) | 11<br>(5, 17) | 74<br>(38, 144) | 72<br>(32, 306) | 1014<br>(194, 2169) | 15.3<br>(2.9, 32.8) | 133<br>(114, 154) |
| *Western Pacific* | 48<br>(42, 55) | 15<br>(7, 24) | 85<br>(39, 174) | 140<br>(54, 659) | 1496<br>(289, 3010) | 31.2<br>(6.0, 63.4) | 96<br>(83, 110) |
| Income Level |  |  |  |  |  |  |  |
| *Low income* | 40<br>(30, 52) | 2<br>(1, 3) | 91<br>(48, 164) | 113<br>(48, 484) | 44<br>(-21, 155) | 1.1<br>(-0.5, 3.7) | 80<br>(59, 103) |
| *Lower middle-income* | 137<br>(114, 163) | 20<br>(9, 32) | 75<br>(39, 141) | 81<br>(34, 342) | 1544<br>(243, 3477) | 11.3<br>(1.7, 25.4) | 275<br>(228, 326) |
| *Upper middle-income* | 89<br>(76, 104) | 34<br>(16, 53) | 67<br>(28, 143) | 102<br>(38, 465) | 3476<br>(755, 6807) | 39.0<br>(8.5, 77.2) | 179<br>(152, 209) |

**Figure 1:**
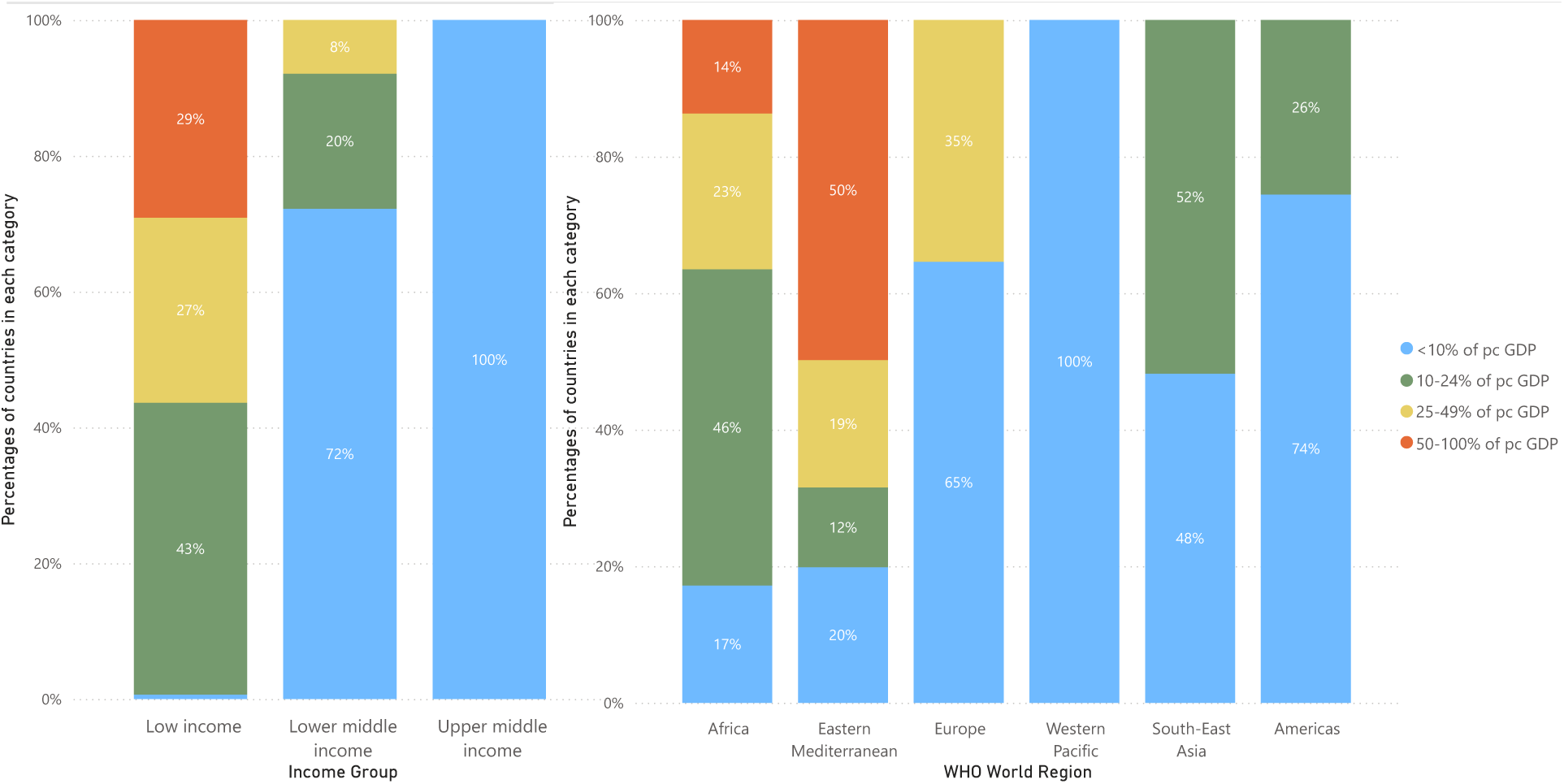
Distribution of estimated cost per DALY averted for individual countries, as a fraction of country per capita GDP. ICER = incremental cost-effectiveness ratio, DALY = disability-adjusted life year, pc GDP = per capita Gross Domestic Product.

### Additional economic outcomes

Aggregated across all countries, net monetary benefits (monetary value of health improvements minus costs) were $5.1 (1.0, 10.4) billion, the return on investment was 19.1 (3.8, 38.5), and the cost-savings produced by adopting a low-dose approach (as compared to 1500mg daily calcium supplementation) were $534 (440, 637) million. Table 2 reports economic outcomes aggregated by WHO region and income level. Figure 2 shows how the return on investment is distributed geographically.

**Figure 2:**
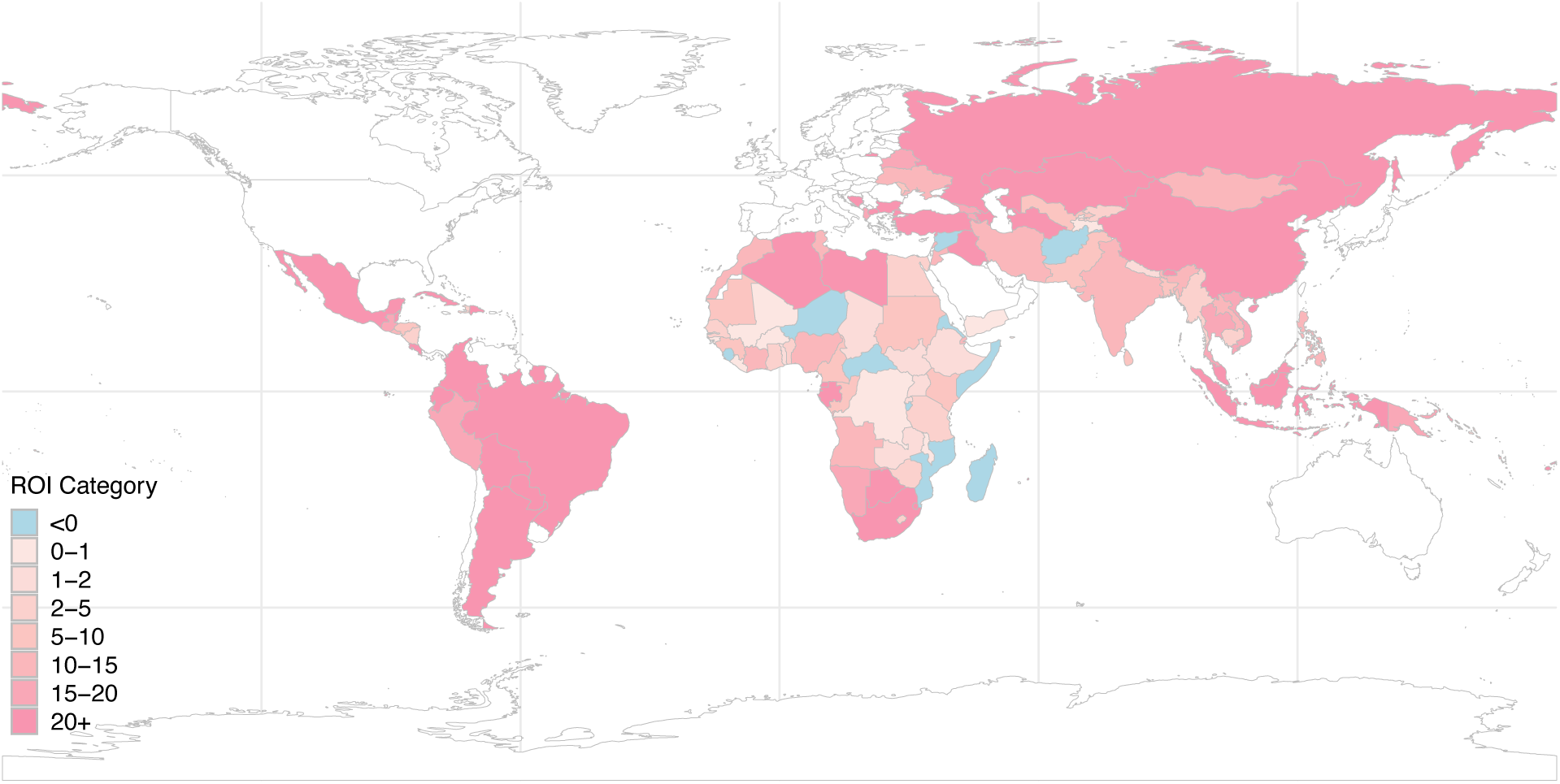
Return on investment by country. Results exclude high-income countries, countries not assessed as having low dietary calcium among women of reproductive age, or countries with insufficient data.

### Sensitivity analyses

Figure 3 and Table S5 show how changes in individual parameters affect the ICER and the NMB of the intervention across all LMICs, as estimated through the one-way sensitivity analyses. For both outcomes, the most influential parameter was the risk ratio for preterm birth, which varied between 0.60 and 0.97. Even with the higher of these values, the cost-effectiveness was still less than $500 per DALY averted. Other influential parameters were the proportion of births that were preterm, intervention adherence, and the cost-effectiveness threshold.

**Figure 3:**
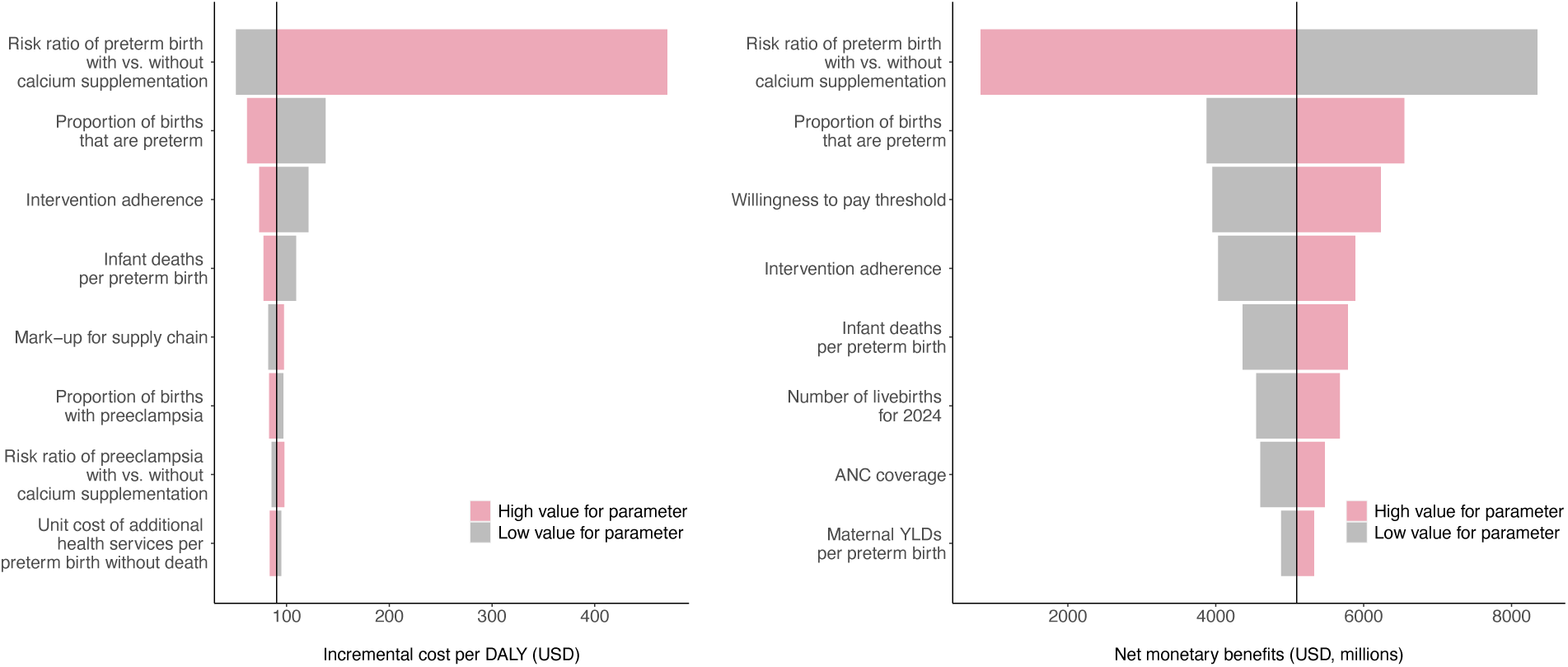
Sensitivity of cost-effectiveness ratio and net monetary benefits for all modeled countries to changes in key input parameters. ANC = antenatal care, YLD = years lived with disability, USD = 2022 US dollars, DALY = disability-adjusted life year.

Tables S6 and S7 present health impact, cost, and cost-effectiveness results aggregated across all LMIC under several alternative analytic assumptions. Compared to the main analysis, scenarios assuming high adherence for all intervention recipients increased DALYs averted by 49% (8.8 million DALYs averted). Scenarios assuming high adherence and full coverage increased DALYs averted by 147% (14.7 million DALYs averted). A scenario assuming that low-adherence intervention recipients would receive 50% of the intervention effect increased DALYs averted by 24% (7.4 million DALYs averted) and reduced the ICER by 24% ($69 per DALY averted). Using an alternative source for the ratios for intervention efficacy reduced DALYs averted by 25% (4.4 million DALYs averted) and increased the ICER by 36% ($123 per DALY averted). When we excluded the cost-savings from averted care for pregnancy complications this increased the ICER by 26% ($114 per DALY averted). Figure S3 compares intervention costs and health outcomes to cost-effectiveness thresholds for this conservative scenario.

Finally, using a calcium unit cost of $0.015 reduced the cost per DALY averted by 31% ($62 per DALY averted), while a calcium unit cost of $0.03 increased the cost per DALY averted by 62% ($147 per DALY averted), as compared to the main analysis.

## Discussion

This study assessed the cost-effectiveness of low-dose (500mg/day) calcium supplementation during pregnancy to prevent preeclampsia and preterm birth in LMIC populations with low dietary calcium intake. Findings show that, for the 2024 birth cohort, low-dose calcium supplementation during pregnancy would be cost-effective for preventing preeclampsia and preterm birth in 119 of 129 modeled LMICs, costing US$90 per DALY averted across all countries. Specifically, low-dose calcium supplementation intervention would prevent 1.3 million preterm births, 1.8 million preeclampsia cases, and 5.9 million DALYs, for a cost of $267 million. In addition, the intervention would save $56 million across these countries. These results show that low-dose calcium supplementation would be highly cost-effective in the large majority of LMICs, a finding that did not change across the different sensitivity analyses and analytic assumptions we tested.

In this analysis, the majority of estimated health benefits resulted from reducing infant deaths due to preterm birth, consistent with the current high burden of mortality from this cause,^2^ and the magnitude of health benefits were larger for countries with a high burden of infant deaths from prematurity. Health benefits were also greater in countries with higher ANC coverage.

When compared to country-specific cost-effectiveness thresholds, the relative cost-effectiveness of the intervention was greatest in countries with higher per-capita income. In several of these countries, the cost-savings from averted care for pregnancy complications were estimated to be greater than intervention costs. Even in a conservative analysis that excluded these cost-savings, cost-effectiveness ratios were well below the suggested cost-effectiveness thresholds for all middle-income countries.^21^ The ten countries in which the intervention was not found to be cost-effective represent some of the poorest countries in the analysis, for which any new intervention would need an extremely low cost-effectiveness ratio to be considered cost-effective. The return on investment results showed a similar pattern (higher ROI values estimated for higher income countries), but even when aggregated across all countries the return on investment (19.1) puts this intervention among the best investments for improving health.^9,23^ These favorable cost-effectiveness results (and high ROI estimate) are notable given the conservative approach taken to valuing health gains. This approach, based on the opportunity cost of healthcare spending,^21^ results in more stringent cost-effectiveness thresholds and lower ROI estimates than approaches based on individual willingness-to-pay for health gains, or historical cost-effectiveness thresholds indexed at 1x and 3x per capita GDP.^24^

This study represents the first cost-effectiveness evaluation of low-dose calcium supplementation during pregnancy. Several studies have evaluated the cost-effectiveness of calcium supplementation for pregnant women, focusing on the 1000-1500mg daily dosage levels included in WHO guidelines.^6–9^ Unsurprisingly—given the substantial cost-savings estimated for low-dose compared to high-dose supplementation—the cost-effectiveness ratios estimated in this analysis are generally lower than those estimated in past analyses. Similarly, the cost-effectiveness of low-dose calcium supplementation appears favorable when compared to other interventions focused on addressing preeclampsia and preterm birth,^25–27^ though less cost-effective than low-dose aspirin for the prevention of preterm birth^28^ and dexamethasone for managing preterm birth.^29^ Importantly, these interventions are not mutually-exclusive, and the best approach to preventing preeclampsia and preterm birth may involve a combination of interventions.

The results of this analysis showed substantial uncertainty, particularly health impact estimates. The primary source of this uncertainty was the risk ratio of preterm birth with calcium supplementation. Other major sources of uncertainty were the base rate of preterm birth and intervention adherence. Estimates of the cost-savings from averted care for pregnancy complications also showed wide uncertainty intervals. While these uncertainties did not affect the cost-effectiveness conclusions, they indicate the potential value of additional research on intervention impacts. Moreover, substantial between-trial heterogeneity has been observed for the effect of calcium supplementation on preeclampsia and preterm birth. An ongoing individual patient data meta-analysis may explain this heterogeneity and elucidate individual-and population-level factors that modify the impact of calcium supplementation on maternal and infant outcomes,^30^ which could be used to refine future economic evaluations. In particular, it is unclear whether parity is an effect modifier of the risk reduction produced by calcium supplementation. This is important because many trials are conducted among nulliparous women, yet preeclampsia risks differ according to the number of prior births.^31^ Evidence on the effect of parity on calcium’s prevention benefits is needed. The sensitivity analyses also demonstrate how low ANC coverage in some countries, and sub-optimal adherence, limit potential intervention impact. Research on innovative intervention modalities to maximize access and adherence would be valuable. The magnitude of several cost components (calcium unit costs, wastage rates, supply chain costs) will become clearer if and when routine supplementation is introduced, allowing cost-efefctiveness estimates to be refined.

Additional limitations of this analysis include the need to impute data for many countries, and the decision to ignore stillbirth and multiple births when calculating birth outcomes. The different definitions used for preeclampsia (which is typically defined narrowly within trials, but more broadly in clinical practice) are an additional source of uncertainty. A potentially larger limitation is the exclusion of recipient costs and cost-savings from the costing perspective. The economic burden of pregnancy complications falls on families as well as the health system,^32,33^ and these costs would be included if the economic analysis had adopted a societal perspective. However, there was insufficient evidence to include these recipient costs in the analysis. Their inclusion would likely have made the intervention appear more attractive. Similarly, we did not assess the long-term intervention consequences for cognitive development and productivity, however, a prior study has demonstrated these benefits to be substantial.^34^ While we only considered LMIC in this analysis, the cost-effectiveness of calcium supplementation (and the comparative advantages of low-dose supplementation) is also relevant for high-income countries. Research focused on these settings is needed.

In summary, this study found that low-dose calcium supplementation provided during pregnancy could produce major health benefits in all settings considered, would be cost-saving compared to the currently recommended calcium dosage, and would be cost-effective in the large majority of countries. In addition to necessary evidence on intervention effects, values, equity, acceptability and feasibility, these findings suggest low-dose calcium supplementation should be considered for inclusion in the WHO-recommended package of interventions for routine antenatal care.

## Declaration of interests

The authors declare no conflicts of interest.

## Contributors statement

HPS, PD, AM, AVK, VS, MMS, RP, HMM, CRS, ABP, WWF and NAM conceptualized the study. HPS, PD, HC, AM, VSP, RP, CY and NAM planned the analysis. HC undertook the analysis. HPS, PD, HC, AM, AVK, VS, MMS, RP, CY, HMM, CRS, ABP, WWF and NAM reviewed and interpreted results. HPS, PR and NAM drafted the manuscript. HC, AM, AVK, VS, MMS, RP, CY, HMM, CRS, ABP, and WWF edited the manuscript.

NAM and HC have verified the data. All authors were responsible for the decision to publish.

## Data Availability

Analytic code and data inputs are available from a publicly-accessible repository (https://zenodo.org/).

## Acknowledgements

This study was funded by the Bill & Melinda Gates Foundation (grant numbers: OPP1172660 and INV-049467). We thank Nandita Perumal, Ulla Griffiths, and Elias Katani for input on data sources and analytic approaches of this study.

## Data sharing statements

Analytic code and data inputs to be submitted to a publicly-accessible repository (https://zenodo.org/) once finalized following journal review.

## Notes

### Competing Interest Statement

The authors have declared no competing interest.

